# Digital technologies for monitoring and improving treatment adherence in children and adolescents with asthma: A scoping review of randomised controlled trials

**DOI:** 10.1101/2021.02.13.21251692

**Authors:** Madison Milne-Ives, Edward Meinert

## Abstract

**Background:** Inadequate paediatric asthma care has resulted in potentially avoidable unplanned hospital admissions and morbidity. A wide variety of digital technologies have been developed to help monitor and support treatment adherence for children and adolescents with asthma. However, existing reviews need to be updated and expanded to provide an overview of the current state of research around these technologies and how they are being integrated into existing healthcare services and care pathways.

**Objective:** The purpose of this scoping review is to provide an overview of the current research landscape and knowledge gaps regarding the use of digital technologies to support the care of children and adolescents with asthma.

**Methods:** The review was structured according to the Preferred Reporting Items for Systematic Reviews and Meta-Analyses extension for Scoping Reviews (PRISMA-ScR) and Population, Intervention, Comparator, Outcome, and Study (PICOS) frameworks. Five databases (PubMed, the Cochrane Central Register of Controlled Trials (CENTRAL), Web of Science, EMBASE, and PsycINFO) were systematically searched for studies published in English from 2014 on. One reviewer screened references, selected studies for inclusion based on the eligibility criteria, and extracted the data, which were synthesised in a descriptive analysis.

**Results:** A wide variety in study characteristics - including the number and age of participants, study duration, and type of digital intervention - was identified. There was mixed evidence for the effectiveness of the interventions; 6 of the 9 studies that evaluated treatment adherence found improvements, but the evidence was inconsistent for asthma control (4/9 found no evidence of effectiveness, and only one found significant evidence) and health outcome variables (4/7 found no evidence of effectiveness). The 5 studies that examined patient perceptions and assessments of acceptability and usability had generally positive findings.

**Conclusions:** Despite the range of different digital interventions being developed to support the monitoring and treatment adherence of children and adolescents with asthma, there is limited evidence to suggest that they achieve their range of intended outcomes. Stronger evidence of their effectiveness at achieving their specific aims is needed, as this will support decisions and research about their cost-effectiveness and how these technologies can best integrate with existing clinical care pathways. This research is necessary to determine which interventions are worth supporting and adopting in the clinical care pathways.

## Introduction

### Background

Globally, and in the UK, asthma is the most common chronic illness affecting children [1–3] and can have serious health consequences. It is one of the key causes of urgent hospital admissions and morbidity in children [3,4]. This is a particularly urgent problem in the UK: out of all the Organisation for Economic Co-operation and Development (OECD) countries, the UK has the third highest risk of death due to paediatric asthma [3]. Asthma-related hospital admissions contribute to the economic burden on the UK healthcare system [5]. It has been estimated that up to two thirds of asthma-related hospital admissions could potentially be avoided through improvements to preventative care systems [4]. The variation of mortality across countries suggests that many of the negative outcomes of childhood asthma - for patients and healthcare systems - are potentially avoidable [4,5]. National reviews have supported the need to improve the management of paediatric healthcare in the UK, stating that the care provided for children and young people with asthma was insufficient [6].

A large, and growing, number of digital technologies have been developed to help support the care and self-management of people with asthma [7–9]. Previous systematic reviews have found that digital interventions can help support asthma health management, particularly by improving medication adherence [10,11]. However, other reviews have identified mixed results in terms of effectiveness (depending on the outcome examined) [9] and app quality [8]. Reviews have also identified limitations in the literature such as inadequate descriptions of the digital interventions, a lack of economic analyses, and small sample sizes [10,12].

To be effective, patients need to be willing to use these digital interventions. While digital interventions have been shown to be generally acceptable to the wider population [11], special consideration is needed when evaluating digital interventions for children and young people. Adolescents are a particularly challenging group to treat, as they are at risk of not attending appointments and falling through a gap if there is not an adequate transition from paediatric to adult healthcare services [2]. Poor health literacy and self-management skills also affect adolescents’ adherence and health outcomes [2]. Attitudes towards electronic monitoring devices were found to be mixed in adolescents, depending on how they perceived the intervention. Among those who viewed asthma as a serious threat, the monitoring device was viewed as reassuring. However, many adolescents were suspicious of the device, reporting concerns that it would get them in trouble if they didn’t not adhere properly to their medication and beliefs that their healthcare providers did not trust them to take the medication [13]. This demonstrates the need to examine digital interventions tailored specifically at children and young people, as their needs and responses to the interventions may not be the same as the general population.

### Rationale

There are several systematic reviews that examined various topics related to digital interventions for asthma management. However, none of the reviews provide a comprehensive and current overview of the field. Some of these reviews examine a general population, including both children and adults [9,11,12], while those reviews that limit the scope to children and young people focus on particular technologies or outcomes [10,14,15]. Additionally, no reviews were found that examined how the technologies are integrated into current clinical care pathways for children and adolescents with asthma.

A comprehensive review of systematic reviews was conducted that captured a large number of outcomes, including cost effectiveness [12]. However, this review was published in 2014, and given the rapid evolution of digital technology [16], the state of the field has changed since the review was conducted; for instance, electronic inhaler monitoring is a relatively new development [17,18], with smart inhalers only recently becoming commercially available [19]. Unni et al.’s review [9] did analyse studies of children and adults separately, but only 16 studies concerning children were included. The review does include a wide range of outcomes - adherence, health outcomes, and user perceptions - but only PubMed and EMBASE databases were searched for the study, which raises the concern that some relevant studies might have been missed [9].

Other systematic reviews have focused specifically on children and young people. However, they limited the scope either with respect to outcome (e.g. a focus on treatment adherence [14]) or type of digital technology (e.g. only mobile apps [10] or smart devices [15]). Several searches of keywords (“asthma” and “child” OR “paediatric” OR “pediatric” and “digital OR technology OR mHealth OR eHealth”) on PROSPERO only identified one relevant registration, which was planned, but not executed, by academics associated with the current research team [20].

Beyond supporting patients’ self-management of their asthma, digital technologies can bridge an information gap that affects the quality of asthma care. By collecting data about symptoms, medication adherence, and other relevant factors, digital technologies can provide healthcare professionals with a large body of information that enables them to personalise asthma care plans and focus on preventative measures [21]. A small study of American physicians found that, while they recognized the potential benefits of integrating digital technologies in asthma care for adolescents, they also noted a number of barriers and concerns [22]. However, more research is needed to understand how digital interventions are currently integrating with healthcare services [21].

Given the extent and variety of the literature on technological interventions to support asthma care in children and young people, there is a need for a scoping review to provide an overview of the state of the literature and to identify knowledge gaps [23]. An overview of the different types of digital technologies and the different ways they are being integrated with healthcare systems will help to inform the development of effective, technologically-enhanced care pathways for children with asthma.

### Objectives and Research Questions

The primary objectives of the scoping review were to assess and summarize the current state of the literature on digital technology-supported asthma care for young people and to identify any gaps. Three research questions were developed to focus the review:

1. How is research on technologically-supported asthma pathways being designed and conducted?
2. What is known about their effectiveness of digital technologies to support treatment adherence and remote symptom monitoring in children and adolescents?
3. What is the state of knowledge about integrating technology into clinical care pathways for paediatric asthma?

## Methods

The review was structured following the Preferred Reporting Items for Systematic Reviews and Meta-Analyses Extension for Scoping Reviews (PRISMA-ScR; Appendix A) [24] and the search strategy was developed using the Population, Intervention, Comparator, Outcome, and Studies (PICOS) framework (see Table 1). A preliminary review of the literature was used to extract MeSH terms and keywords for the search. The search was performed in five databases using the University of Plymouth’s search tool Primo - PubMed, the Cochrane Central Register of Controlled Trials (CENTRAL), Web of Science, EMBASE, and PsycINFO - with slightly adjusted search terms to fit the specific structure of each database (see Appendix B). The search terms were grouped into four themes - asthma, asthma management, children, and digital technology - that were joined with the structure: asthma (MeSH OR Keywords) AND asthma management (MeSH OR Keywords) AND children (MeSH OR Keywords) AND digital technology (MeSH OR Keywords). A complete record of the specific search terms and strings used for each database and the number of references returned is provided in Appendix B. The database searches were completed on 30 December 2020, except for the CENTRAL database, which was searched on 31 December 2020.

**Table 1.** PICOS framework

|  |  |
| --- | --- |
| Population | Children and young people under 18 years old with asthma |
| Intervention | Any digital health technology aiming to support monitoring or treatment adherence of children and adolescents with asthma |
| Comparator | No comparator is required |
| Outcome | The primary outcome was the evidence for the digital interventions at improving monitoring or treatment adherence. Secondary outcomes included how the research was conducted, evidence for improved health outcomes, cost-effectiveness, and integration of the technology with healthcare systems. |
| Study types | Randomised controlled trials that evaluate at least one digital technology to support the care of children with asthma. |

### Inclusion criteria

The review included studies that evaluate digital technologies that aim to support the monitoring or treatment adherence of children and adolescents with asthma. Digital technologies included, but were not limited to, mobile or web applications, smart devices, and other phone or internet-based interventions. Initially, randomised controlled trials (RCTs), quantitative, qualitative, cohort, and case study types were eligible for inclusion. Given the number of studies identified, only RCTs were included in the review.

### Exclusion criteria

Studies of patients over the age of 18 were excluded, there were three exceptions that reached full-text review and had both child and adult participants that were included in the review. Studies that were published before 2014 were excluded because, due to the rapid evolution of digital technology [16], earlier studies do not necessarily reflect the current state of the field. Studies that merely describe an intervention without evaluating it were excluded. Studies that are published in languages other than English were also excluded, as the review team did not have the necessary resources to assess them.

### Screening and Article Selection

References were exported to the citation management software EndNote X9 for storage and duplicate removal. Due to the large number of references returned, an initial screening was conducted by inputting keywords relating to the inclusion and exclusion criteria into the EndNote X9 search function. This was done in several stages, due to the limitation of the number of search criteria that could be specified at once (see Appendix C for details). Searches of keywords to exclude were based on common features of irrelevant studies that were identified in a manual search. The remaining titles and abstracts were screened by the reviewer (with articles excluded with reasons), and final eligibility was determined by full-text reviews of the remaining references.

### Data Extraction

Outcomes were extracted by the reviewer from a full-text review of all of the included articles into a table structured according to the three research questions (see Appendix D). Key outcomes were pre-determined based on a preliminary review of the literature; however, because of the expected variety of reported outcomes, relevant outcomes that were not pre-specified in the PICOS or data extractions tables were also considered for inclusion in the final review.

### Data Analysis and Synthesis

The data extracted from the studies about the key outcomes listed in Table 2 were assessed using a descriptive analysis and summarised to provide an overview of the state of the literature. For the outcomes relating to effectiveness, the number of studies that found strong evidence of effectiveness was compared to the number of studies that assessed that outcome to provide a synthesis of the state of the evidence for that outcome. Implications of the findings were examined in the discussion.

**Table 2.** Article information and data extraction

| Article information | Data to be extracted |
| --- | --- |
| General study information |  |
|  | Year of publication |
|  | Sample size |
|  | Age of participants |
| Digital technology |  |
|  | Type of digital technology |
|  | Healthcare setting used in |
| Evaluation |  |
|  | Effect of technology on behavioural outcomes (e.g. medication adherence, symptom monitoring and reporting, etc.) |
|  | Effect of technology on health outcomes |
|  | Cost-effectiveness of the intervention |
|  | Integration of the technology with a health system / care pathway |
|  | Participant perceptions |
|  | Acceptability |
|  | Usability |
|  | Other key performance indicators reported |

## Results

### Included Studies

6,314 articles were retrieved from the search of the five databases (see Appendix B). 1,029 duplicates were removed by the EndNote X9 software, and a further 5,193 were screened out using keyword searches in EndNote (see Appendix C). The titles and abstracts of 92 studies were screened and articles were excluded with reasons. Of these articles, 25 were selected for the full-text review, and 20 were selected for inclusion in the review. Six of the references referred to one study and were either conference abstracts or did not include the final results of the randomised controlled trial. The paper with published results of the RCT of that study was identified and included [25]. The reasons for exclusion in the full-text review stage are detailed in Figure 1.

**Figure 1.**
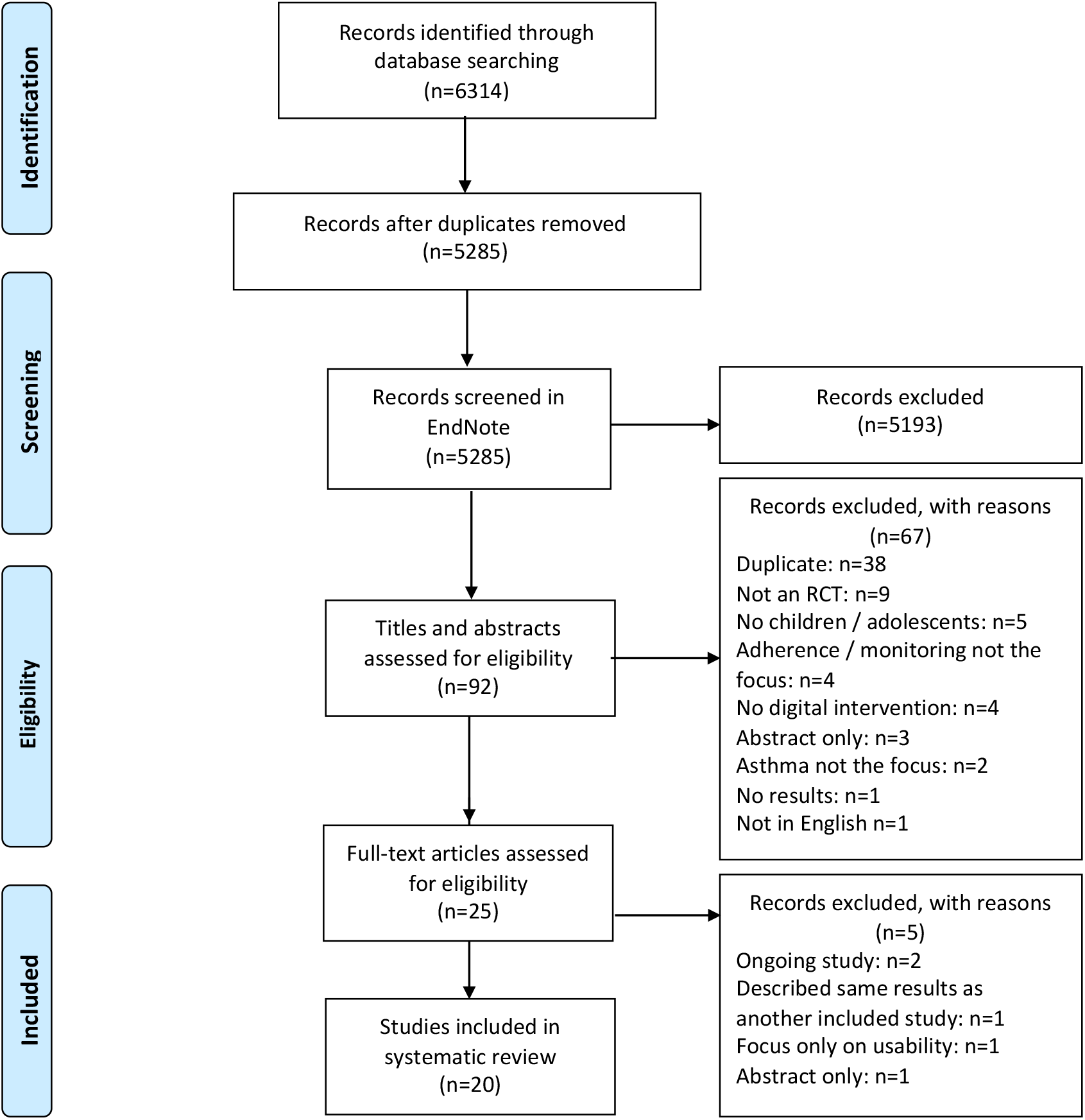
Preferred Reporting Items for Systematic Reviews and Meta-Analyses (PRISMA) flow diagram

### Study Characteristics

All of the studies included in the review were randomised controlled trials (RCTs), and were limited to those that included monitoring or adherence functions and aims. Despite these restrictions to the scope of the review, the included studies had a wide variety of study durations, sample sizes, age ranges, and types of digital intervention. Over a third of the references identified as eligible during title and abstract screening only had abstracts available (7/20) [26– 32]. They were included in the analysis, using whatever data was provided; two of the abstracts only presented interim results [26,27]. Four studies were analysed by nine separate articles and abstracts: the ADolescent Adherence Patient Tool (ADAPT) study [25,33,34], a study comparing web-ACT and FENO monitoring with standard care [35,36], a study of inhaler electronic monitoring devices (EMDs) with audiovisual reminders [37,38], and a study of a real-time medication monitoring (RTMM) device with SMS reminders [28,39].

There was a wide range in study durations, from 3 weeks [40] to 24 months [41], with the most common length of follow-up being 6 or 12 months (n=4 [25,31,33,34,37,38,42] and n=3 studies [27,28,35,36,39] for each). There was also a large variety of numbers of participants included in the 15 studies, ranging from 22 [30] to almost 1200 [41], with an average of approximately 230 participants and a median of 209 [28,39].

There were no distinctive categories of ages that emerged from the studies. Of the 15 distinct studies, only two used the same age range (4-11 years old [28,39,43]). Three studies included adult participants as well as child or adolescent participants [26,29,42]. The youngest participants included in a study were 3 years old [41]. Of the studies that focused on participants under 18, the span of ages eligible for inclusion in each study ranged from 6 years (ages 12-17 [40]) to 15 years (ages 4-18 [35,36]).

4 studies took place across multiple centres [26,28,32,35,36,39], and most of the rest were associated with large medical centres [40–43] or clinics [27,31]. The remaining 5 studies recruited from or were associated with a hospital emergency department [37,38], community pharmacies [25,33,34], Howard University [29], and impoverished, rural school districts [44]. One study did not specify the healthcare setting of the study [30].

### Types of Digital Interventions

Several different types of digital interventions for monitoring and/or improving medication adherence were examined in the studies included in this review. The most common type of intervention, evaluated by a third of the studies (5/15), was EMDs. However, these EMDs all varied on their features, which included: audiovisual reminders [37,38], text messages [28,39], alarms [27], and app or online sources that could be synced to provide personal feedback [26,27], educational content [26], reminders [31], and capture adherence data [31].

Apps were another common intervention evaluated; three studies specifically evaluated 3 different app-based interventions. These included the ADolescent Adherence Patient Tool (ADAPT) app that connects adolescents to their community pharmacist through a desktop application and enables them to monitor symptoms and adherence, chat with peers and their pharmacist, watch short educational movies, and set medication alarms [25,33,34]. Another app, CHANGE Asthma, was developed for children by five pediatricians and modified based on feedback from a pilot of 24 caregivers. It uses short videos and games and an asthma action plan to improve asthma knowledge and control [43]. The third app evaluated (AsthmaWin) also included an asthma action plan, but focused more on monitoring symptoms and medication adherence [29].

Other types of interventions evaluated included web-based monitoring programs [35,36] (one of which was a component of a Virtual Asthma Clinic [32]), a speech recognition automated telephone program to improve medication adherence [41], text message medication reminders [42], a website and text-based reminder system (MyMediHealth) [40], a remote directly observed therapy (R-DOT) tool to improve inhaler use and adherence [30], and a school-based educational telemedicine intervention that provided interactive video sessions for children, caregivers, and school nurses [44].

### Evidence of Effectiveness

A number of different outcome measures were used by the studies to evaluate the interventions, but results about effectiveness were inconsistent. Outcome with the highest proportion of studies finding a significant, positive effect was for improving medication adherence. The reported effectiveness of interventions and improving asthma control and health outcomes was very mixed. Patient feedback regarding acceptability and usability was generally high.

### Treatment or Medication Adherence

9 studies evaluated the effectiveness of their intervention at improving treatment or medication adherence. Two thirds (6/9) reported significantly higher adherence in the intervention group compared to the control group [25,28,31,37,39–41]. Of the remaining three studies, two reported higher adherence in the intervention group compared to the control group, but no analysis of significance was provided [27,44]. The final study, which evaluated a SMS reminder system, found a decline in adherence over the intervention and control periods in both groups [42].

3 of the 6 studies that found a significant difference in adherence between groups evaluated EMDs [28,31,37,39]. The others evaluated the speech recognition automated telephone program [41], the website and text-based reminder system (MyMediHealth) [40], and the ADAPT app [25].

Only one study each evaluated the effectiveness of improving inhaler use and symptom monitoring, both of which found improvements. Shields et al. found that R-DOT improved inhaler technique equally in immediate and delayed intervention groups [30]. Perry et al. found significantly higher self-reports of peak flow meter use in the intervention group compared to the control group [44].

### Asthma Control and Healthcare Visits

There were very mixed results in the nine studies that evaluated asthma control as an outcome. Four of the nine studies found either no effect of the intervention on asthma control [25,26,39] or no significant difference between groups [43]. However, Real et al. did find an significant positive association between degree of app use and asthma control [43].

Another four studies reported improved asthma control in the intervention group compared to the control group [27,29,30,32], although only one of these studies demonstrated statistical significance [32]. The final study found that asthma control could be maintained after a clinically relevant reduction in inhaled corticosteroids in the web-based monitoring condition [35,36].

Only two studies evaluated the effect of the intervention on healthcare visits, but neither found any differences [32,41].

### Health and Quality of Life Outcomes

The overall effect of digital interventions on health outcomes is also unclear. Four of the seven studies that evaluated health outcomes (such as quality of life or symptom free days) found no significant improvement [25,28,35,36,39,44]. However, the other three studies reported significant improvements in self-reported quality of life [40], asthma morbidity scores [37], and number of symptom free days [32].

### Patient Perceptions, Acceptability, and Usability

Five studies examined outcomes related to patient perceptions, acceptability, or usability. These all reported generally high satisfaction and acceptability [33,34,38,40] or a desire to continue using the intervention [29,31].

### Cost-effectiveness

Only one study (two articles) explicitly assessed cost-effectiveness [28,39]. The authors found that costs were higher in the intervention group, and although this difference was not statistically significant [39], the technology was deemed not cost-effective because it was not associated with significant improvements in health outcomes [28].

### Integration with clinical care pathways

Most of the studies included in the review (9/15, or 11 of the 20 articles) did not explicitly discuss how the digital intervention they were evaluating was integrated with clinical care pathways [27–31,37–40,42,43]. There were few studies that described sending data from the interventions back to physicians to update the patients’ health records or inform care, although this potential would likely be feasible for many of them. For the few that did, integration of the intervention with the healthcare system was generally reported positively.

Even among those that did describe a specific link between the intervention and the healthcare system, the specific details about integration were not a primary focus of the paper. For instance, some of the studies that monitored symptoms or adherence produced treatment advice based on data analysis from the system algorithms [35,36], sent physicians warnings if a patient was out of a certain threshold [26]. The Virtual Asthma Clinic, which also sent feedback to physicians if a patient’s asthma control scores were low, was found to be successful at increasing asthma control and symptom-free days and was proposed by the authors as a partial replacement for outpatient visits [32]. Details of how these systems were integrated with the healthcare system were not described.

One study whose intervention was significantly integrated with the healthcare system was the ADAPT app study [25,33,34]. One of the aims of the intervention was to increase collaboration and communication between adolescents and pharmacists because of the increasing role of pharmacists as healthcare providers in the Netherlands [25]. Pharmacists involved in the intervention reported valuing the improved contact with patients and found the intervention satisfactory, useful in fulfilling their role, and not time-consuming [34]. This was in contrast to the perceptions of pharmacists who did not participate in the intervention, who identified time constraints as a barrier to the use of mHealth [34]. However, a barrier was identified because the ADAPT app’s ‘stand-alone’ desktop interface for pharmacists was not integrated with the pharmacy’s general information system [34]. This study highlights the potential value of deliberate and considered efforts to integrate new digital health technologies for asthma management with existing health systems.

The speech recognition telemedicine intervention was another study that demonstrated integration with the healthcare system: the telemedicine system was integrated with the hospital’s electronic health record (EHR; EpicCare) to provide personalised calls to patients and is compatible with all standard EHR systems [41].

The attempt of one study [44] to involve primary care providers in the intervention was not successful. Treatment prompts with medication recommendations based on caregiver reports and guidelines were provided to the participants’ primary care providers. These were found to be ineffective; of the 141 prompts sent out for individual participants, the request for feedback received a response from only one primary care provider [44].

## Discussion

### Summary of Findings

There was a lot of variety in the studies examined in this review; study duration ranged from 3 weeks to 2 years, the number of participants ranged from 22 of 1187, and - although the review was focused on children and adolescents - there was a wide range of ages studied, with no distinct age groups emerging from the studies. There were also several different types of digital interventions analysed in the RCTs, with electronic monitoring devices and mobile apps the most common. However, the integration of these technologies with existing clinical care pathways and health systems was not extensively discussed in the majority of studies.

The review found inconsistent evidence for the effectiveness of the digital technologies at achieving their various aims. The most support was found for the effectiveness of the interventions at improving treatment or medication adherence (6/9 studies found evidence of effectiveness). The results of studies assessing the impact of the intervention on asthma control and health outcomes were mixed, with some studies reporting positive effects and others no significant effect. Across the studies, evaluations of patient perceptions, acceptability, and usability were generally positive.

### Limitations

One limitation of this review is that the screening, article selection, and data analysis were only conducted by one researcher. Although the PRISMA-ScR framework was used to ensure the review performed and reported the necessary components of a scoping review, the lack of validation from another, independent reviewer means that there is a greater potential for bias to have been introduced in the selection or analysis of the studies.

Given time constraints when conducting the analysis, the data was extracted according to research questions (see Appendix D), with data for specific outcomes being identified from that table. In addition, a risk of bias assessment was not performed on the studies. While this is not a standard requirement for scoping reviews, it is a limitation of the study, as it would have contributed to the assessment of the first research question by providing an analysis of the quality of the research being conducted on technologically-supported asthma pathways.

Another limitation is that the research questions and aims were adjusted after the search had been performed. They were changed before any screening or selection took place, but may have resulted in relevant articles being missed because the search terms were established for a slightly different scope. Because of time limitations, no hand searches of the references of reviews retrieved in the initial search were performed, which also could have resulted in eligible articles being overlooked.

### Meaning and Future Research

The large number of studies identified in the initial search and the variety of technological interventions to support paediatric asthma care demonstrate the broad scope of this research area. This review identified few strong trends with regards to how technologically-supported asthma pathways for children and young people are being researched. A theoretical framework for determining what ages to study or how to stratify children and young people into age groups might be useful for future research by enabling meta-analyses to be conducted. Currently, there is no consensus in the literature on how to group children of various ages for research.

This review found that there are a wide variety of different digital interventions being explored. However, strong evidence of their effectiveness at achieving various aims is still lacking. Notably, there was almost no consideration of the cost-effectiveness of the intervention in the studies examined. This will be a key area for future evaluations of these technologies to consider, so that limited healthcare resources can be deployed to create the greatest value [45].

Another key area for future research will be around the integration of these digital solutions into clinical pathways. As with cost-effectiveness, this review found that the majority of studies did not explicitly consider or evaluate how the technology they were examining would interact with existing health systems. The potential benefit of integrating patient-reported data with patients’ health records to inform care plans and pathways is likely feasible for many of the technologies assessed but was not examined as a key outcome of the technology. Acceptability and usability data likewise focused primarily on patient users. Understanding how these technologies can best support and interact with existing clinical pathways could help to inform their design, improvement, and sustainable adoption.

## Conclusion

The purpose of this scoping review was to examine and summarise the state of the literature on technologically-enhanced asthma care pathways for children and young people. A large body of research is ongoing in this area and spans a wide range of technologies and ages. Although there was some evidence found for their effectiveness - particularly for improving treatment and medication adherence - further research is needed to establish the effectiveness of the interventions at improving asthma control and other health outcomes. There was little research that described or assessed the integration of these technologies with existing clinical care pathways or the cost-effectiveness of the interventions. These are key areas for future research to examine so that the value for patients and healthcare systems of the variety of digital technologies currently being developed can be comprehensively evaluated and compared.

## Data Availability

This is a review of secondary literature and all data is accessible via public databases.

## Author Contributions

IW conceived the key research questions. JG and KH developed and submitted the previous PROSPERO registration that was used as the basis for the protocol. The scoping review was executed and drafted by MMI with revisions from EM and IW.

**Appendices**

## Appendix A. PRISMA-ScR Checklist

**Preferred Reporting Items for Systematic reviews and Meta-Analyses extension for Scoping Reviews (PRISMA-ScR) Checklist**

| SECTION | ITEM | PRISMA-ScR CHECKLIST ITEM | REPORTED ON PAGE # |
| --- | --- | --- | --- |
| **TITLE** |  |  |  |
| Title | 1 | Identify the report as a scoping review. | Title Page |
| **ABSTRACT** |  |  |  |
| Structured summary | 2 | Provide a structured summary that includes (as applicable): background, objectives, eligibility criteria, sources of evidence, charting methods, results, and conclusions that relate to the review questions and objectives. | 1 |
| **INTRODUCTION** |  |  |  |
| Rationale | 3 | Describe the rationale for the review in the context of what is already known. Explain why the review questions/objectives lend themselves to a scoping review approach. | 3 |
| Objectives | 4 | Provide an explicit statement of the questions and objectives being addressed with reference to their key elements (e.g., population or participants, concepts, and context) or other relevant key elements used to conceptualize the review questions and/or objectives. | 4 |
| **METHODS** |  |  |  |
| Protocol and registration | 5 | Indicate whether a review protocol exists; state if and where it can be accessed (e.g., a Web address); and if available, provide registration information, including the registration number. | N/A |
| Eligibility criteria | 6 | Specify characteristics of the sources of evidence used as eligibility criteria (e.g., years considered, language, and publication status), and provide a rationale. | 4-5 |
| Information sources* | 7 | Describe all information sources in the search (e.g., databases with dates of coverage and contact with authors to identify additional sources), as well as the date the most recent search was executed. | 4 |
| Search | 8 | Present the full electronic search strategy for at least 1 database, including any limits used, such that it could be repeated. | 18-22 (Appendix B) |
| Selection of sources of evidence† | 9 | State the process for selecting sources of evidence (i.e., screening and eligibility) included in the scoping review. | 5 |
| Data charting process‡ | 10 | Describe the methods of charting data from the included sources of evidence (e.g., calibrated forms or forms that have been tested by the team before their use, and whether data charting was done independently or in duplicate) and any processes for obtaining and confirming data from investigators. | 5 |
| Data items | 11 | List and define all variables for which data were sought and any assumptions and simplifications made. | 6 |
| Critical appraisal of individual sources of evidence§ | 12 | If done, provide a rationale for conducting a critical appraisal of included sources of evidence; describe the methods used and how this information was used in any data synthesis (if appropriate). | N/A |
| Synthesis of results | 13 | Describe the methods of handling and summarizing the data that were charted. | 6 |
| **RESULTS** |  |  |  |
| Selection of sources of evidence | 14 | Give numbers of sources of evidence screened, assessed for eligibility, and included in the review, with reasons for exclusions at each stage, ideally using a flow diagram. | 7 (Figure 1) |
| Characteristics of sources of evidence | 15 | For each source of evidence, present characteristics for which data were charted and provide the citations. | 7-8 |
| Critical appraisal within sources of evidence | 16 | If done, present data on critical appraisal of included sources of evidence (see item 12). | N/A |
| Results of individual sources of evidence | 17 | For each included source of evidence, present the relevant data that were charted that relate to the review questions and objectives. | 25-34 (Appendix D) |
| Synthesis of results | 18 | Summarize and/or present the charting results as they relate to the review questions and objectives. | 8-10 |
| **DISCUSSION** |  |  |  |
| Summary of evidence | 19 | Summarize the main results (including an overview of concepts, themes, and types of evidence available), link to the review questions and objectives, and consider the relevance to key groups. | 10-11 |
| Limitations | 20 | Discuss the limitations of the scoping review process. | 11 |
| Conclusions | 21 | Provide a general interpretation of the results with respect to the review questions and objectives, as well as potential implications and/or next steps. | 11-12 |
| **FUNDING** |  |  |  |
| Funding | 22 | Describe sources of funding for the included sources of evidence, as well as sources of funding for the scoping review. Describe the role of the funders of the scoping review. | 12 |

JBI = Joanna Briggs Institute; PRISMA-ScR = Preferred Reporting Items for Systematic reviews and Meta-Analyses extension for Scoping Reviews.
\* Where *sources of evidence* (see second footnote) are compiled from, such as bibliographic databases, social media platforms, and Web sites.
† A more inclusive/heterogeneous term used to account for the different types of evidence or data sources (e.g., quantitative and/or qualitative research, expert opinion, and policy documents) that may be eligible in a scoping review as opposed to only studies. This is not to be confused with *information sources* (see first footnote).
‡ The frameworks by Arksey and O'Malley (6) and Levac and colleagues (7) and the JBI guidance (4, 5) refer to the process of data extraction in a scoping review as data charting.
§ The process of systematically examining research evidence to assess its validity, results, and relevance before using it to inform a decision. This term is used for items 12 and 19 instead of "risk of bias" (which is more applicable to systematic reviews of interventions) to include and acknowledge the various sources of evidence that may be used in a scoping review (e.g., quantitative and/or qualitative research, expert opinion, and policy document).
From: Tricco AC, Lillie E, Zarin W, O'Brien KK, Colquhoun H, Levac D, et al. PRISMA Extension for Scoping Reviews (PRISMA-ScR): Checklist and Explanation. *Ann Intern Med.* 2018;169:467–473. doi: [10.7326/M18-0850](https://doi.org/10.7326/M18-0850).

**Appendix B.**
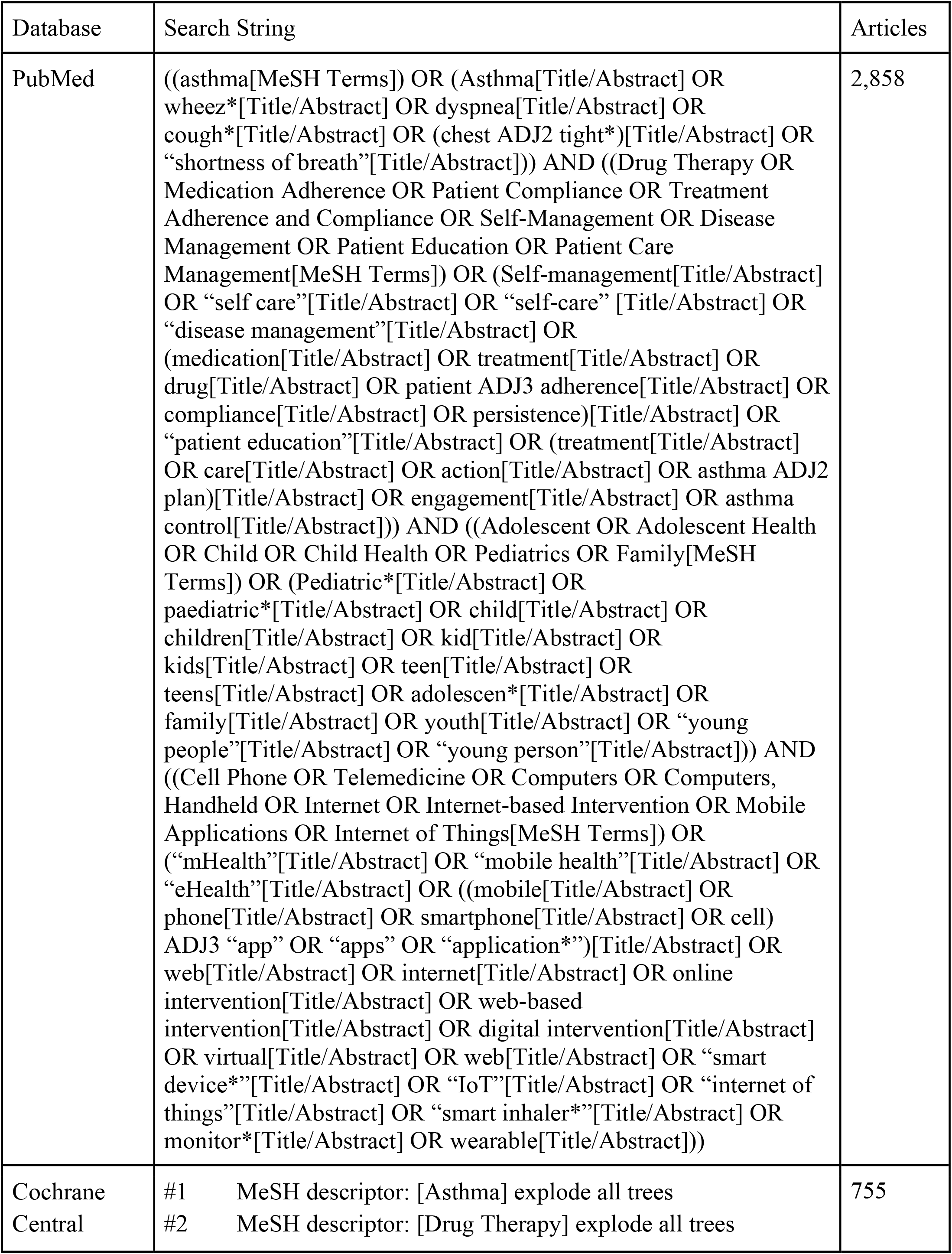
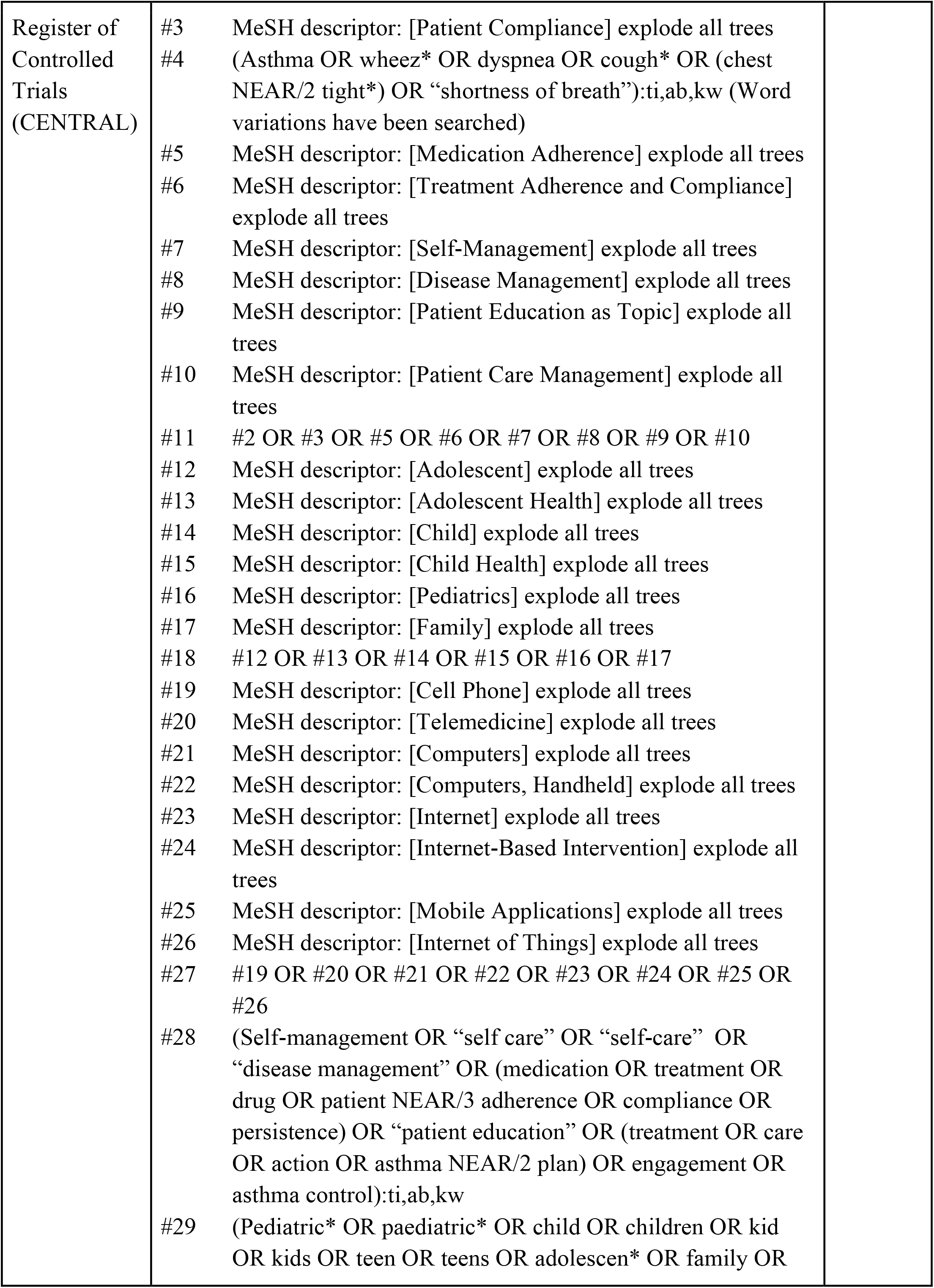
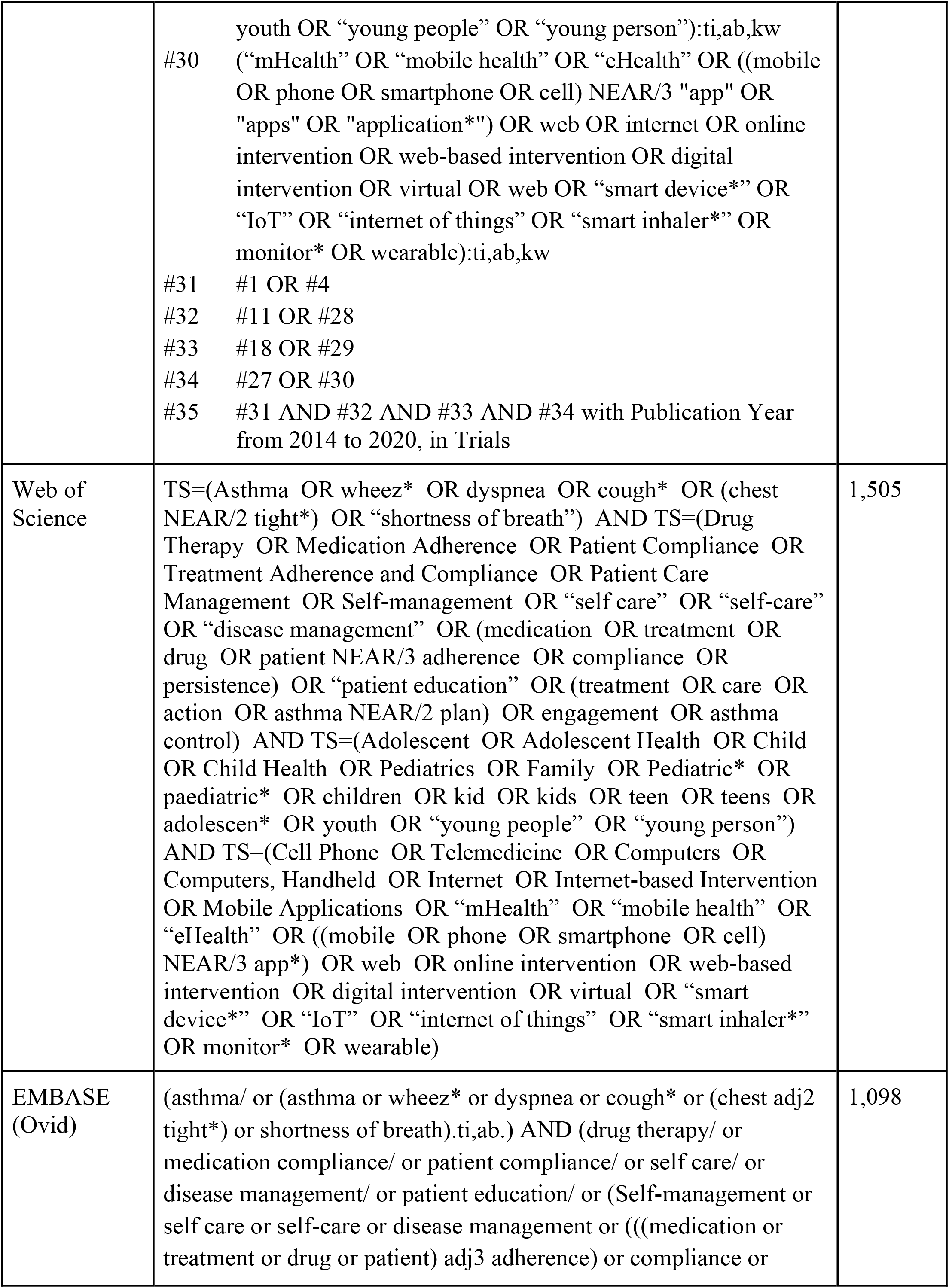
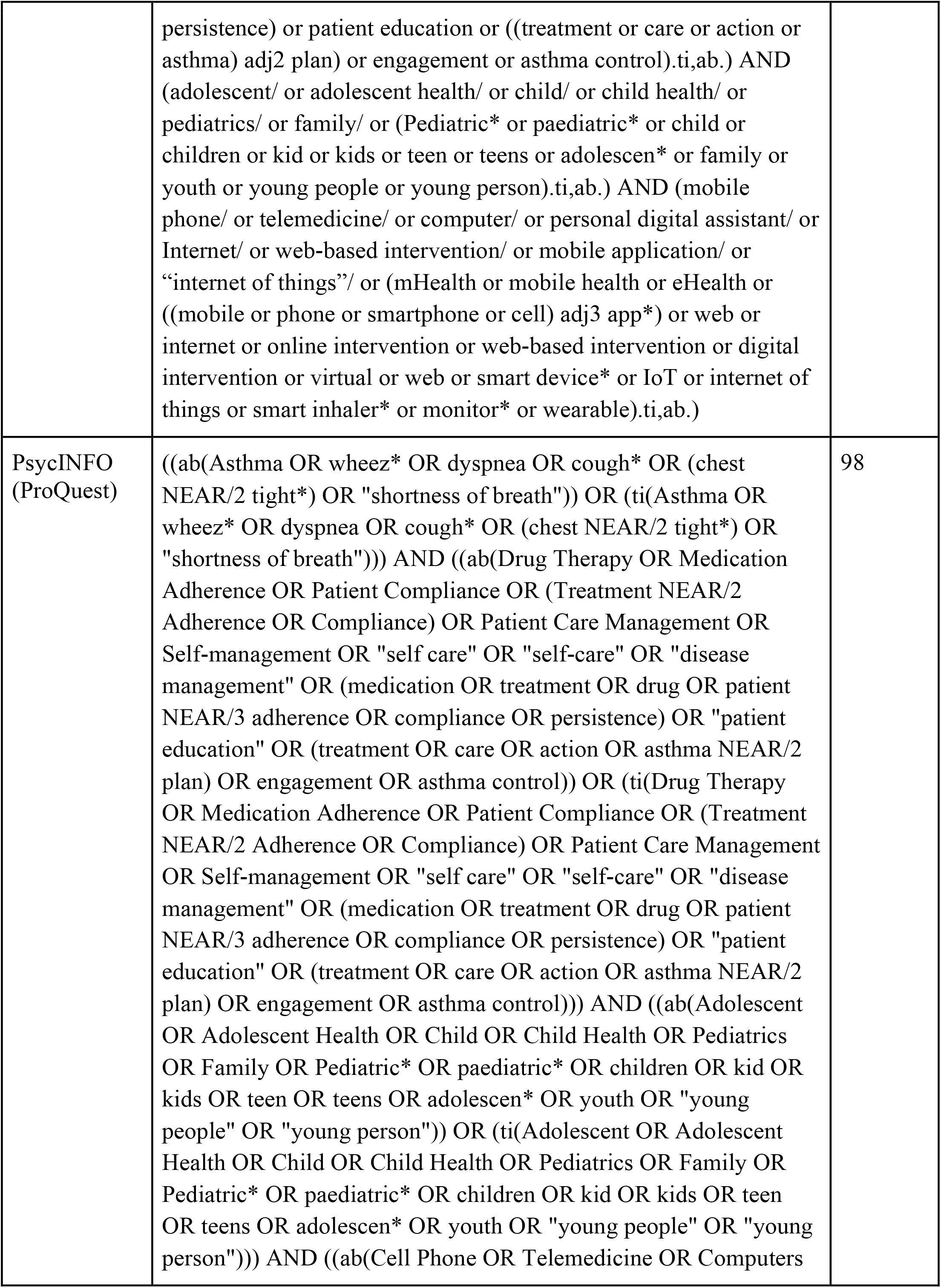
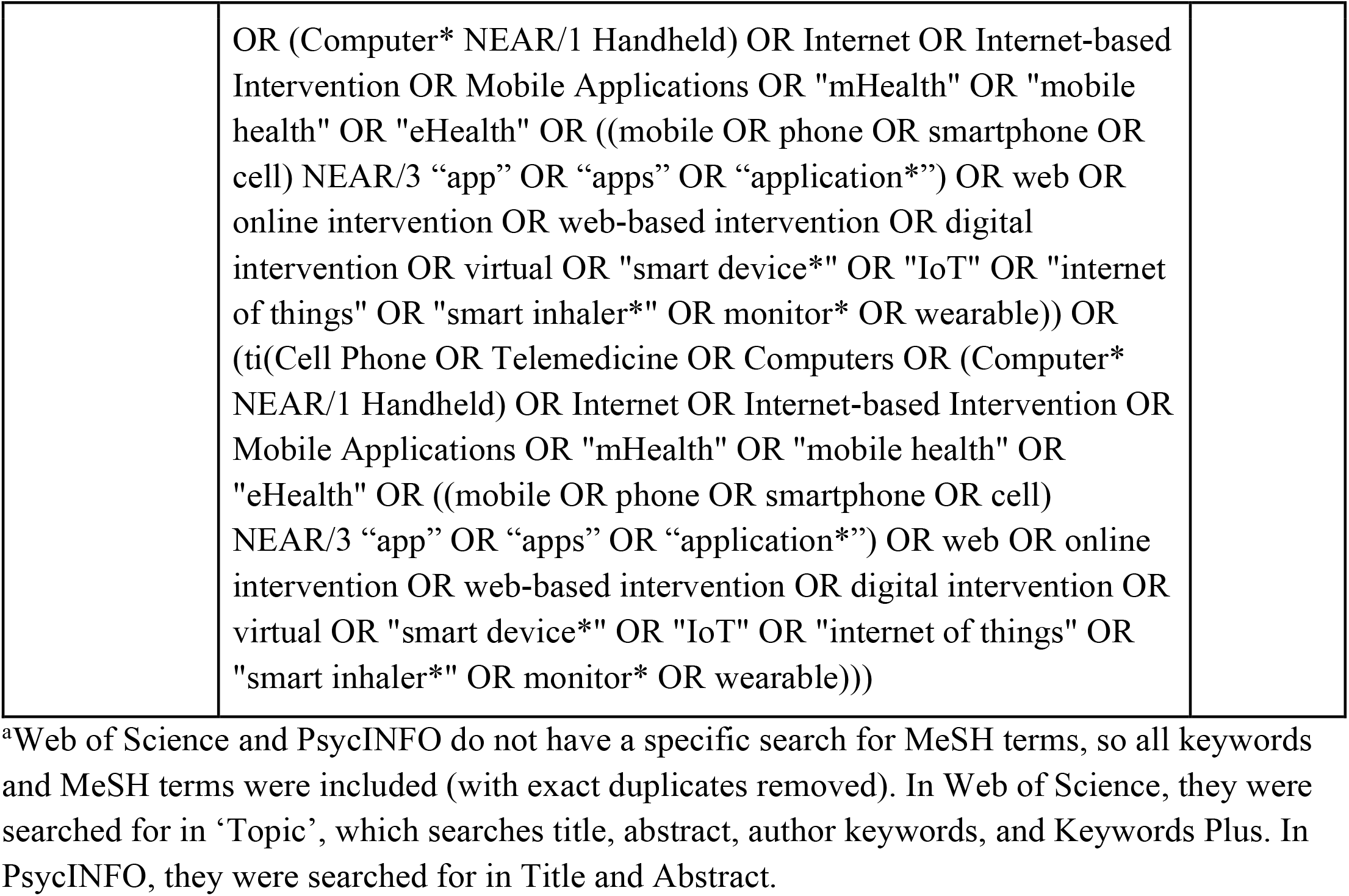
Search record

**Appendix C:**
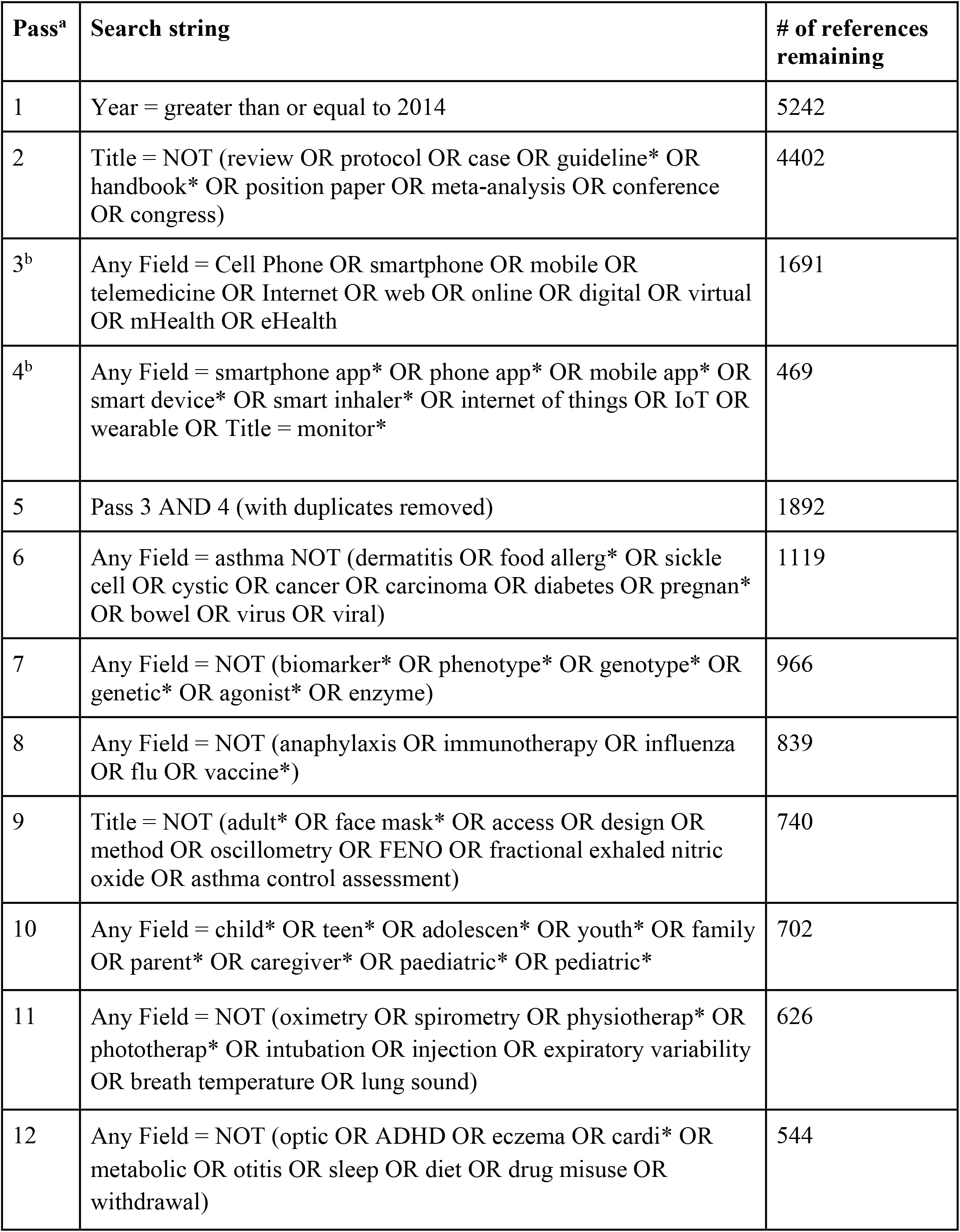
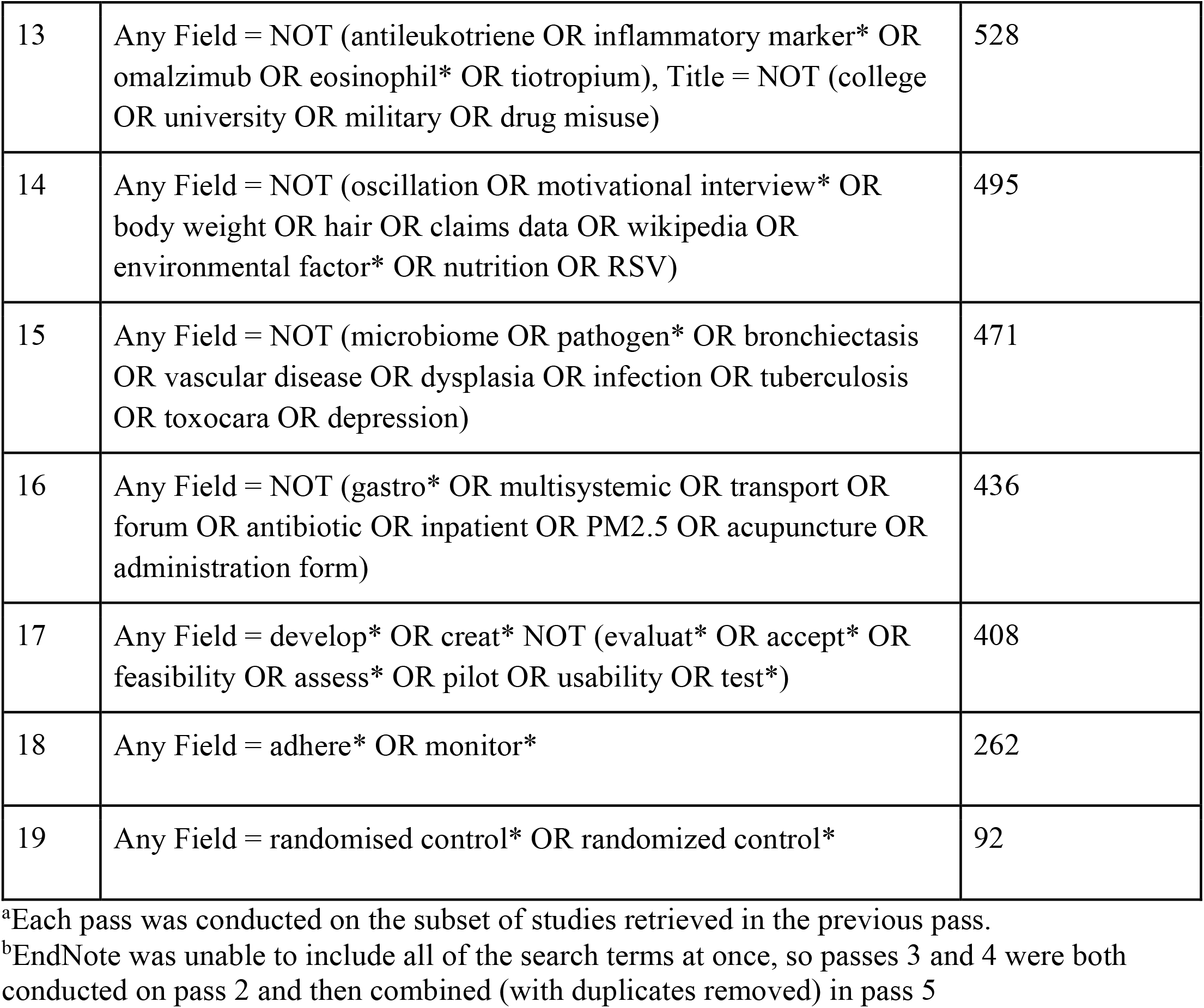
Endnote search criteria

| Pass<sup>a</sup> | Search string | # of references remaining |
| --- | --- | --- |
| 1 | Year = greater than or equal to 2014 | 5242 |
| 2 | Title = NOT (review OR protocol OR case OR guideline* OR handbook* OR position paper OR meta-analysis OR conference OR congress) | 4402 |
| 3<sup>b</sup> | Any Field = Cell Phone OR smartphone OR mobile OR telemedicine OR Internet OR web OR online OR digital OR virtual OR mHealth OR eHealth | 1691 |
| 4<sup>b</sup> | Any Field = smartphone app* OR phone app* OR mobile app* OR smart device* OR smart inhaler* OR internet of things OR IoT OR wearable OR Title = monitor* | 469 |
| 5 | Pass 3 AND 4 (with duplicates removed) | 1892 |
| 6 | Any Field = asthma NOT (dermatitis OR food allerg* OR sickle cell OR cystic OR cancer OR carcinoma OR diabetes OR pregnan* OR bowel OR virus OR viral) | 1119 |
| 7 | Any Field = NOT (biomarker* OR phenotype* OR genotype* OR genetic* OR agonist* OR enzyme) | 966 |
| 8 | Any Field = NOT (anaphylaxis OR immunotherapy OR influenza OR flu OR vaccine*) | 839 |
| 9 | Title = NOT (adult* OR face mask* OR access OR design OR method OR oscillometry OR FENO OR fractional exhaled nitric oxide OR asthma control assessment) | 740 |
| 10 | Any Field = child* OR teen* OR adolescen* OR youth* OR family OR parent* OR caregiver* OR paediatric* OR pediatric* | 702 |
| 11 | Any Field = NOT (oximetry OR spirometry OR physiotherap* OR phototherap* OR intubation OR injection OR expiratory variability OR breath temperature OR lung sound) | 626 |
| 12 | Any Field = NOT (optic OR ADHD OR eczema OR cardi* OR metabolic OR otitis OR sleep OR diet OR drug misuse OR withdrawal) | 544 |
| 13 | Any Field = NOT (antileukotriene OR inflammatory marker* OR omalizumab OR eosinophil* OR tiotropium), Title = NOT (college OR university OR military OR drug misuse) | 528 |
| 14 | Any Field = NOT (oscillation OR motivational interview* OR body weight OR hair OR claims data OR wikipedia OR environmental factor* OR nutrition OR RSV) | 495 |
| 15 | Any Field = NOT (microbiome OR pathogen* OR bronchiectasis OR vascular disease OR dysplasia OR infection OR tuberculosis OR toxocara OR depression) | 471 |
| 16 | Any Field = NOT (gastro* OR multisystemic OR transport OR forum OR antibiotic OR inpatient OR PM2.5 OR acupuncture OR administration form) | 436 |
| 17 | Any Field = develop* OR creat* NOT (evaluat* OR accept* OR feasibility OR assess* OR pilot OR usability OR test*) | 408 |
| 18 | Any Field = adhere* OR monitor* | 262 |
| 19 | Any Field = randomised control* OR randomized control* | 92 |

<sup>a</sup>Each pass was conducted on the subset of studies retrieved in the previous pass.
<sup>b</sup>EndNote was unable to include all of the search terms at once, so passes 3 and 4 were both conducted on pass 2 and then combined (with duplicates removed) in pass 5

**Appendix D.** Data Extraction Table

| Paper | RQ1. How is research on tech-supported asthma pathways being designed and conducted? | RQ 2. What is known about the effectiveness of tech to support treatment adherence and remote symptom monitoring? | RQ 3. What is the state of knowledge about integrating tech into clinical care pathways? |
| --- | --- | --- | --- |
| [35]<br>NOTE: same study as Voorend-van Bergen | - 1 year study, children 4-18, comparing monitoring strategies to adapt care - standard care (ACT once every 4 months) vs monthly web-ACT vs ACT every 4 months + FENO<br>- Recruited by their own paediatrician from general hospitals and tertiary referral centres<br>- Multicentre, prospective, partly blinded, parallel-group, three-arm randomised controlled superiority trial on monitoring strategies in asthmatic children with a follow-up of 1 year | - neither strategy improved symptom free days compared to standard care after 1 year<br>- Monthly web-based ACTs resulted in a clinically relevant decrease of ICS dose, while maintaining asthma control.<br>- FENO group: increase in asthma control in children under 12 | -not specifically discussed<br>- both strategies provided in addition to usual care - integrated? |
| [41] | -24 month study, ages 3-12, computerized speech recognition telephone intervention vs usual care condition<br>- SR program created with parent focus groups, script development, and beta testing<br>- personalised with data from EHR and automated, option to get callback from nurse/pharmacist<br>- study was conducted within Kaiser Permanente Colorado, a large, group-model health maintenance organization | -inhaled corticosteroid adherence was 25.4% higher in the intervention group than in the usual care group (24-month mean [SE] adherence, 44.5% [1.2%] vs 35.5% [1.1%], respectively; $P < .001$)<br>- no differences in healthcare visits<br>- strong potential for low-cost SR adherence programs integrated with an electronic health record | - intervention integrated in addition to usual care<br>- integrated with an electronic health record<br>- more than half of physicians use an EHR and current projections indicate that more than 90% of clinicians will be using an EHR within the decade.<sup>21</sup><br>Further, the intervention in this study leveraged EpicCare, the most widely used ambulatory care EHR in the United States. The capacity to identify and sort patients by EHR indicators in this project is<br>standard in all EHR systems, including eClinicalWorks, McKesson, Cerner, Allscripts, and GE Healthcare |
| [42] | -64 patients age 12 to 22 years with a diagnosis of poorly controlled persistent asthma in a 6-month longitudinal crossover study<br>- Adherence was objectively monitored in 22 of the participants<br>- participants were given the opportunity to personalize their text message reminders. They could schedule non-asthma medication reminders, appointment reminders, or other messages of their choice | - adherence declined in both groups over the intervention and control periods. This suggests that 3 months of messages did not create a long-term habit of ICS use<br>- | - not discussed explicitly<br>- were recruited at clinic visits<br>- should be easy to incorporate alongside standard care |
| [37] | - patients aged 6–15 years who attended the regional emergency department<br>- followed up every 2 months for 6 months<br>- randomly assigned patients to receive an electronic monitoring device for use with their preventer inhaler with the audiovisual reminder functions either enabled to support adherence to inhaled corticosteroids (intervention group) or disabled (control group) | - Median percentage adherence was 84% (10th percentile 54%, 90th percentile 96%) in the intervention group, compared with 30% (8%, 68%) in the control group ($p < 0.0001$)<br>- significant improvements in adherence to inhaled corticosteroids in school-aged children with asthma<br>- change in asthma morbidity score from baseline to 6 months was significantly greater in the intervention group than in the control group ($p = 0.008$) (more reduce in int group) | - Not explicitly discussed |
| [38] | - Children 6 to 15 years presenting with asthma to the hospital emergency department and prescribed inhaled corticosteroids were included<br>- Device quality control tests were conducted. Questionnaires on device acceptability, utility and ergonomics were completed at six months | - Acceptability scores were high, with higher scores in the reminder than non-reminder group (median, 5th-95th percentile: 4.1, 3.1–5.0 versus 3.7, 2.3–4.8; $p < 0.001$).<br>- Most (>90%) rated the device easy to use.<br>- Feedback was positive across five themes: device acceptability, ringtone acceptability, suggestions for improvement, effect on medication use, and effect on asthma control | - Not specifically discussed |
|  |  | - failure rates of 13–16% indicate the importance of quality control |  |
| [28] *NOTE: conference abstract only | - multi-center, randomized controlled trial. Included were 209 children (<12 years) with moderate to severe asthma, followed for 12 months<br>- all received a Real-Time Medication Monitoring system, in intervention group it sent texts to parents whose child appeared to miss a dose | - RTMM increases inhalation adherence, but there is no evidence of better health outcomes in this patient population within the first year. In these circumstances, this is not a cost-effective intervention.<br>- Costs were higher in the intervention group | - Not specifically discussed |
| [40] | - impact of MyMediHealth (MMH) – a website and a short messaging service (SMS)-based reminder system – on medication adherence and perceived self-efficacy in adolescents with asthma<br>- block-randomized controlled study in academic pediatric outpatient settings, 98 adolescents enrolled<br>- MMH users were asked to create a medication schedule and receive SMS reminders at designated medication administration times for 3 weeks | - Compared to controls, we found improvements in self-reported medication adherence ($P = .011$), quality of life ($P = .037$), and self-efficacy ($P = .016$).<br>- Subjects reported high satisfaction with MMH; however, the level of system usage varied widely<br>- significant racial disparity in the rate of MMH adoption | - not specifically discussed |
| [25] | - Cluster randomized controlled trial in 66 Dutch community pharmacies.<br>- Asthma patients aged 12–18 years were invited to participate, based on pharmacy medication refill records.<br>- 234 adolescents (147 in the control group and 87 in the intervention group) completed the study<br>- The main study outcome was self-reported medication adherence, measured with the Medication Adherence Report Scale (MARS).<br>- Secondary outcomes were asthma control and quality of life. Outcomes were measured at start ($t = 0$ months) and at the end of follow-up ($t = 6$ months) | - We did not find an intervention effect in the total study population, i.e., only in non-adherent patients.<br>- there was a positive effect of the intervention on medication adherence (MARS +2.12, $p = 0.04$).<br>- This effect was stronger (MARS +2.52, $p = 0.02$) in poor adherent adolescents with uncontrolled asthma ($n = 74$).<br>- No effect of the intervention was observed on asthma control or quality of life. | - current study used pharmacists as the healthcare provider, because pharmacists are increasingly expected to support appropriate use of medication in integrated care settings<br>- Increased collaborations between pharmacists and physicians may facilitate the identification of uncontrolled patients with low adherence rates. |
|  |  |  | - Pharmacists can subsequently support these patients with their medication use, by implementing mHealth interventions.<br>- Adherence could also be measured by using pharmacy refill records.<br>However our study period covered only six months, and in the Netherlands patients usually collect chronic medication once every three months. |
| [34] *NOTE: same study as above | - explore experiences, barriers, and facilitators of pharmacists and patients towards the use of the interactive ADOlescent Adherence Patient Tool (ADAPT)<br>- the perceptions of pharmacists towards mHealth interventions in general were explored<br>- setting: dutch community pharmacies | - Most patients (78%) would recommend the ADAPT intervention to others, and thought that the pharmacy was the right place for mHealth aiming to support adherence (63%).<br>- The possibility to monitor asthma symptoms was highly appreciated by patients and pharmacists.<br>- Pharmacists were satisfied with ADAPT intervention (96%), and using the intervention was not time consuming (91%).<br>- The ADAPT intervention promoted contact with patients (74%) and facilitated the healthcare providing role of pharmacists (83%). | - Providing extra care for patients was one of the main reasons for using mHealth (by both pharmacist groups).<br>- Pharmacists who delivered the ADAPT intervention valued the improved patient contact<br>- Another important facilitator for further implementation is the integration of mHealth in the pharmacy information system, because a 'stand-alone' desktop program restrained the integration with the<br>pharmacist's workflow<br>- MHealth interventions can facilitate the pharmacist's responsibilities and promote contact with patients. This is important nowadays, because pharmacists are expected to combine their management role with more healthcare providing roles, and there is an ongoing shift towards integrated care settings |
| [33] *NOTE: same study as above | - Explore the use and the effective engagement of adolescents (aged 12 to 18 years) with the Adolescent Adherence Patient Tool (ADAPT)<br>- The ADAPT app was connected to a desktop application of the patient's own community pharmacist<br>- included: questionnaire to monitor symptoms, peer and pharmacist chats, medication alarm, short movies, adherence questions | - 86 adolescents (mean age 15.0, SD 2.0 years) used the ADAPT app 17 times (range 1-113) per person. Females used the app more often than males ($P=.01$) and for a longer period of time ($P=.03$).<br>- The questionnaires to monitor symptoms and adherence were used by most adolescents.<br>- The total app use did not affect adherence; however, activity in the pharmacist chat positively affected medication adherence ($P=.03$), in particular, if patients sent messages to their pharmacist ($P=.01$)<br>- Adolescents have different preferences when using an mHealth app, as there was a wide variety in app usage per person<br>- The questionnaires to monitor asthma symptoms and adherence were used by most adolescents, which provided valuable data for health care providers and<br>patients. Moreover, the use of the pharmacist chat positively affected adherence. |  |
| [26] *NOTE: abstract only, interim results | - intervention: inhaler sensors, personalised feedback and educational content (apps and online interfaces)<br>- control group had sensors only<br>- children and adults (5-80)<br>- 490 enrolled, 368 active/completed at 16 months | - sig diff in rescue inhaler use between groups after 100 days (fewer for int group)<br>- | - physicians could monitor intervention group patients and received proactive warnings if they fall below a threshold |
| [27] *NOTE: abstract only, interim results | - complex intervention comprising electronic adherence monitoring with feedback and reminder alarms can improve clinical outcomes and adherence in childhood asthma<br>- 90 participants were recruited, 25 have completed study, and 60% of follow-up visits have been completed<br>- Children with asthma in the UK were recruited and followed up in standard clinics for 12 months | - At 12 months, the mean adherence in the intervention arm is currently 83%, compared to 34% in the control arm<br>- The mean number of oral steroid courses in the 12 months is 1.7 in the intervention group and 2.7 in the control group | - not specifically discussed |
| [44] | - cluster randomized trial with rural children, ages 7–14 years (393 enrolled), comparing a school-based telemedicine asthma education intervention to usual care<br>- Each student participated in five 30–45 minute age-appropriate asthma education sessions via telemedicine (live interactive video)<br>- Telemedicine sessions were also conducted for intervention parents/caregivers and school nurses. Each 60–90 minute session was conducted at the school (local community center on weekend/nights). Caregivers participated in 2 sessions and school nurses participated in 1 session.<br>- Telemonitoring sessions were completed prior to the first child education session and at 3 months. During the session, intervention participants described symptoms for the preceding 2 weeks and completed the PedsQL 3.0 survey | - At the end of the intervention, there were no statistically significant differences in reported symptom free days (primary outcome) for either the intervention or usual care group.<br>- Participants in the intervention group reported significantly higher utilization of peak flow meters to monitor asthma and reported taking their asthma medications as prescribed more frequently when compared to the usual care group.<br>- There were no changes in other outcome measures including quality of life, self-efficacy, asthma knowledge, or lung function between groups<br>- Although there was some evidence of behavior change among intervention participants, these changes were inadequate to | - The Primary Care Provider of each intervention participant received a prompt with guidelines-based asthma management<sup>19</sup> at baseline and 3 months. Caregivers and school nurses received copies of the prompt to reinforce recommendations. The prompt included: 1) a summary of education sessions, 2) blank AAP with completion instructions, 3) synopsis of |
|  |  | overcome the significant morbidity experienced by this highly symptomatic rural, impoverished population. | caregiver-reported symptoms and medications and 4) treatment recommendations according to guidelines.<sup>19</sup> For example, if a participant's caregiver reported uncontrolled, recommendations for initiation or step-up of controller therapy were given.<sup>19</sup> PCPs received a 7-question survey to confirm receipt of the prompt, verify accuracy of asthma severity and control assessment. We provided a self-addressed stamped envelope to return the survey and phone/fax/mailling contacts for the research team to answer any questions or to receive additional feedback.<br>- Simply providing a treatment prompt to PCPs with medication recommendations was proven to be ineffective. |
| [43] | - developed and implemented a smartphone application (app) leveraging gamified features entitled CHANGE Asthma ("Clinic, Home, And on the Go Education for Asthma"). We subsequently assessed its impact on asthma control. | - The control and intervention groups both included 20 caregivers with 75% of participants completing follow-up.<br>- Although C-ACT scores among intervention participants | - not discussed |
|  | - Patients aged 4–11 years with a previously documented childhood asthma control test (C-ACT) score of <20, indicating poor control, were recruited<br>- App usage was monitored for 4 months<br>- curriculum iteratively developed by 5 pediatricians and 2 app developers<br>- piloted with caregivers and modified accordingly | significantly improved at follow-up, compared to their own baseline ($P = 0.04$), the change of C-ACT score did not significantly differ from that of the control group ($P = 0.78$).<br>- Among the intervention participants, there was a positive, dose-dependent relationship between app usage time and positive change in C-ACT score ($P = 0.03$). |  |
| [29] *NOTE: abstract only | - AsthmaWin is an iPhone app, developed by CooperSoft Inc. that incorporates a physician-generated asthma action plan & involves daily recording of: (a) peak flow measurements, (b) medication usage—documented with a self-photo, (c) daily symptoms with an automatic reminder to take medications and that can reward completion of data, is intended to enhance patient physician communication<br>- whether the use of this app would improve medication adherence & improve asthma self-management as measured by Asthma Control Test (ACT) scores and peak expiratory flow rate (PEFR) measurements<br>- Forty eight asthmatics, predominantly African American, 13-60 years old were recruited from the Howard University Faculty Practice Plan<br>- | - Documented daily use of the Asthma Action plan leads to improvement in ACTs. PEFRs and medication adherence. Focus groups indicated patient willingness to use the app on an ongoing basis to assist in asthma management and an unwillingness to continue paper journaling.<br>- Reported controller medication usage was 86% among app users and 90% in the paper group. ACT scores in the app group improved by a mean of 3.8 points and by 2.4 points in the paper group. PEFRs improved an average of 9.09% in the app group and 7.82% in the paper group. | - not discussed, although enhanced communication was an aim |
| [30]*NOTE: abstract only | - determine if remote directly observed therapy (R-DOT) could monitor adherence and inhaler technique in children with Difficult to Treat Asthma (DTA)<br>- pilot study: 22 children (aged <15 yrs) with DTA were randomised to either immediate R-DOT for 6 weeks or delayed entry (R-DOT after 6 weeks) and asthma control was assessed at 12 weeks.<br>- mobile phone platform was used to record and send a short video clip (timed and dated, twice a day) of children using their inhaler. The videos were reviewed by a nurse and if needed the child/parent was provided with reminders/re-instruction | - Despite all children being able to demonstrate good inhaler technique at study entry, 80% were deemed not to have good inhaler technique while using R-DOT at home during the first week.<br>- By the end of the 3rd week after nurse led re-instructions all children had good inhaler technique. Both groups improved equally.<br>- R-DOT can test for and improve inhaler technique and monitor adherence and can be used to ensure that a period of<br>optimised therapy has been delivered in DTA. The approach is not device specific and can be used with any type of inhaled therapy.<br>- Engagement with the process was generally good although cases of non-adherence with video uploads included living with a different parent at weekends, life too busy and mobile phone memory issues | - not discussed in abstract, but intervention required nurse involvement |
| [31] *NOTE: abstract only | - 8-17 years, 43 enrolled<br>- Breathe Smart EMD, one each for rescue and controller inhalers<br>- synced with mobile app, sends reminders and captures adherence data | - adherence based on pharmacy records after 3 months was significantly greater in int group than control, equal to mean adherence captured by EMD<br>- 83% desired to continue using EMD<br>- however, adherence in both groups still low | - not discussed |
| [32]<br>*Abstract only | - multicentre, randomised controlled trial with a 16-month follow-up, 210 asthmatic children (6–16 years) treated in eight Dutch hospitals were randomised to usual care (4-monthly outpatient visits) and online care using a virtual asthma clinic (VAC) (8-monthly outpatient visits with monthly web-based monitoring)<br>- In between VAC visits, asthma control was monitored online, filling in a (C-)ACT monthly.<br>- If the (C-)ACT score was $\geq 20$, automatic default messages were emailed with positive and encouraging content.<br>- If the (C-)ACT score was $< 20$, feedback to the participants included advice to check their medication use, an individual action plan and a request to contact their asthma team when symptoms persisted.<br>- In addition, feedback was sent to the asthma team with the request to contact the participant within 2 working days to address the clinical condition of that moment. | - After follow-up, symptom-free days differed statistically between the usual care and VAC groups (difference of 1.23 days, 95% CI 0.42–2.04; $p=0.003$) in favour of the VAC. In terms of asthma control, the Childhood Asthma Control Test improved more in the VAC group (difference of 1.17 points, 95% CI 0.09–2.25; $p=0.03$). No differences were found for other outcome measures. | - Routine outpatient visits can partly be replaced by monitoring asthmatic children via eHealth.<br>- physicians contacted if issues identified<br>- demonstrates that eHealth using the VAC can substitute 50% of routine outpatient visits in paediatric asthma care while the number of SFDs improved significantly after a 16-month follow-up |
| [39] *NOTE same as Goosens study | - multicentre, randomised controlled trial, 209 children (aged 4–11 years) using ICS were recruited from five outpatient clinics and were given an real-time medication monitoring (RTMM) device for 12 months<br>- The intervention group also received tailored SMS reminders, sent only when a dose was at risk of omission | - Mean adherence was higher in the intervention group: 69.3% versus 57.3% (difference 12.0%, 95% CI 6.7%–17.7%).<br>- No differences were found for asthma control, quality of life or asthma exacerbations.<br>- Costs were higher in the intervention group, but this difference was not statistically significant | - not discussed |
| [36] | - multicentre trial with a 1-year follow-up, children aged 4–18 years with a doctor's diagnosis of asthma treated in seven hospitals were randomised to one of the three groups. In the web group, treatment was adapted according to ACT obtained via a website at 1-month intervals; in the FENO group according to ACT and FENO, and in the SC group according to the ACT at 4-monthly visits<br>- 280 included, 268 completed | - change from baseline in symptom-free days did not differ between monitoring strategies. With web-based ACT monitoring, ICS could be reduced substantially while control was maintained. | - treatment was adapted monthly according to the web-based ACT score, while in the FENO group, treatment was adapted to FENO and ACT score at clinic visits every 4 months<br>- The treating physicians were blinded to randomisation group, FENO and ACT. The local investigators, unblinded to ACT and FENO, provided the physicians with treatment advice based on the study algorithms and on the treatment plan<br>- Physicians could deviate from this advice for documented clinical reasons only. |

